# Analytical sensitivity analysis and clinical impact modeling of Rapigen rapid diagnostics tests for malaria

**DOI:** 10.1101/2023.08.14.23292196

**Authors:** Allison Golden, Hannah C. Slater, Ihn Kyung Jang, Sayali Walke, Thanh T. Phan, Greg T. Bizilj, Andy Rashid, Becky Barney, Gonzalo J Domingo

## Abstract

Analytical performance analysis through laboratory benchmarking can more objectively compare the performance of malaria rapid diagnostic tests (RDTs). We present the analytical detection limits of the Rapigen BIOCREDIT Malaria Ag Pf/Pv (pLDH/pLDH), the Rapigen BIOCREDIT Malaria Ag Pf (pLDH/HRPII), and two best-in-class World Health Organization (WHO)-prequalified comparator RDTs, generated using standardized panels containing recombinant antigen, in vitro cultured parasites, international standards, and clinical samples. Detection limits of HRP2, PfLDH, and PvLDH were determined for the Rapigen and comparator RDTs as antigen concentration and in international units (IU)/mL. The Rapigen Ag Pf (pLDH/HRPII) detected 3.9 and 3.9 IU/mL for PfLDH and HRP2, respectively, while the Ag Pf/Pv (pLDH/pLDH) detected 3.9 and 5.0 IU/mL for PfLDH and PvLDH, respectively. The comparator HRP2/PfLDH and HRP2/PvLDH detected 15.6 and 31.3 IU/mL for HRP2 and PfLDH and 15.6 and 50.0 IU/mL for HRP2 and PvLDH, respectively. The RDT clinical sensitivity was predicted through application of analytical detection limits to antigen concentration distributions from clinical symptomatic and asymptomatic cases. Febrile cases would be detected in majority by both standard and Rapigen RDTs, though with increases in the Rapigen RDTs that may be important for clinical cases currently missed by microscopy. Rapigen RDTs were predicted to increase the detection of asymptomatic cases and improve the detection of hrp2 deletions through PfLDH detection. Through the benchmarking and simulation of clinical sensitivity, a method for rapidly assessing the ability of new RTDs to meet clinical needs using high-sensitivity antigen distribution data is presented.

## Introduction

Rapid diagnostic tests (RDTs), typically in the form of lateral flow cassettes, are the primary means of diagnosing malaria, alongside microscopy. RDTs fulfill the need of clinics to have a fast and cost-effective method of diagnosis in the absence of an expert microscopist.

RDTs function by detecting protein antigens, produced by the malaria parasite, which are circulating in the peripheral blood. RDTs used to diagnose *Plasmodium falciparum* (Pf) malaria in the vast majority detect the Pf-specific histidine-rich protein 2 (HRP2), while RDTs used to diagnose either *Plasmodium vivax* (Pv) or malaria caused by any human *Plasmodium* species target the conserved essential enzyme lactate dehydrogenase (LDH). While HRP2 has been the most sensitive marker to date for Pf infection, there are two main challenges to its ongoing use as the sole biomarker in diagnosis of Pf infection. Following treatment, HRP2 can remain in the circulation and persist for weeks beyond clearance of the parasite, resulting in possible false-positive results in individuals who have recently been treated for malaria.^1–3^ The viability of Pf with *hrp2* and the highly homologous *hrp3* gene deletions and the clinical presentation of individuals with *hrp2/hrp3*-deletion strains have led to false-negative RDT results even in symptomatic cases presenting with high parasite densities.^1^ Such deletions have prompted dedicated World Health Organization (WHO) guidance as well as surveillance strategies to monitor their prevalence.^2,3^ Surveillance and clinical data point to increases of *hrp2/hrp3*-deletion prevalence in many countries.^4^ If significant prevalence of hrp deletions are observed, HRP2-based diagnostics become inappropriate for use. Instead, an RDT that also detects falciparum-specific PfLDH may be required, but observed tradeoffs in sensitivity, as compared to detection of Pf through HRP2, further highlight why advances in LDH sensitivity are needed.^5^

Detection of Pv infection requires *Plasmodium vivax*-specific lactate dehydrogenase (PvLDH). This biomarker has two challenges: the assay binding kinetics for pLDH have been poorer than those of HRP2, and lower parasite densities and circulating parasites—and correspondingly lower blood antigen levels—are common with Pv infections.^6^ As a result, RDTs for diagnosing Pv have poorer sensitivity than HRP2-based tests for Pf. Gains have been made in the limits of detection of HRP2 and, more recently, pLDH.^7,8^ Understanding what such gains mean for clinical performance requires analysis of incremental gains in sensitivity, comparison to clinical data, and understanding of the distributions of antigen levels.

A benchmarking protocol was developed to characterize the analytical sensitivity of malaria RDTs for Pf and Pv. The benchmarking panel consists of well-characterized protein antigen sources such as recombinant protein, diluted clinical samples, and cultured parasites in which LDH and HRP2 have been quantified. This benchmarking protocol was applied to WHO-prequalified RDTs as well as two RDTs produced by Rapigen: one RDT for Pf, BIOCREDIT Malaria Ag Pf (pLDH/HRPII) with test lines to detect HRP2 and PfLDH; and one for Pf and Pv, the BIOCREDIT Malaria Ag Pf/Pv (pLDH/pLDH) with test lines to detect PfLDH and PvLDH. The analytical performance is described, and their clinical performance is predicted through modeling.

## METHODS

### Benchmarking panels materials

Whole blood units taken in dipotassium ethylenediaminetetraacetic acid–anticoagulant (K_2_ EDTA) from five healthy donors were used for dilution of panel members (Interstate Blood Bank, Memphis, TN, USA). The individual negative samples were evaluated using the Q-Plex™ Human Malaria (5-Plex) (Quansys, Logan, UT, USA) to confirm that they did not have detectable malaria antigen. The five units were then combined in equal volumes to prepare a pooled negative blood sample.

Recombinant HRP2 tagged with glutathione-S-transferase protein was purchased from Microcoat Biotechnologie (Starnberger See, Germany). Recombinant PfLDH and PvLDH proteins were purchased from MyBioSource (San Diego, CA, USA). Human recombinant LDH was acquired from the University of Queensland Protein Expression Facility (St. Lucia, Australia). The WHO international standards for Pf (product code 16/376) and Pv (product code 19/116) antigens were purchased from the National Institute for Biological Standards and Control (NIBSC) (Hertfordshire, UK). Pf W2 (hrp2+/hrp3+), D10 (hrp2–/hrp3+), HB3 (hrp2+/hrp3–), and Dd2 (hrp2–/hrp3+) strains were obtained from the Biodefense and Emerging Infections Research Resources Repository (Manassas, VA, USA) and the 3BD5 (hrp2–/hrp3–) strain was obtained from the National Institute of Allergy and Infectious Diseases (Bethesda, Maryland, USA). *P. knowlesi* strain A1–H2 was a gift from Dr. Rob Moon (London School of Hygiene and Tropical Medicine, UK). Clinical *P. falciparum*- and *P. vivax*-positive samples used to prepare panels were obtained from Discovery Life Sciences (Santa Barbara, CA, USA).

### Recombinant protein panel members and international standards

Recombinant HRP2-GST and pLDH stock solution concentrations were confirmed by amino acid sequencing (University of Nebraska Medical Center Protein Structure Core Facility, Omaha, NE, USA). Proteins were diluted serially into pooled negative whole blood and aliquoted for storage at −80°C. NIBSC standards 16/376 and 19/116 were reconstituted according to instructions and then serially diluted into the pooled negative whole blood and aliquoted for storage at −80°C.

### Culture panel members

*P. falciparum* strains ITG, Dd2, D10, HB3, and 3BD5 and *P. knowlesi* were prepared at PATH using in vitro culture and synchronization methods previously described.^9–13^ Parasite count was determined via staining of smear and 100x oil immersion objective light microscopy. Parasitized red blood cells were washed with phosphate-buffered saline and pelleted and frozen for long-term storage at −80°C until serially diluted for use in panels. The Pf culture panel containing cultured W2 was purchased from ZeptoMetrix (product code KZMC043, ZeptoMetrix, Franklin, MA).

### Clinical dilution panel members

All clinical positive samples from Discovery Life Sciences used in panels were confirmed positive for *Plasmodium* monoinfection by photo-induced electron transfer polymerase chain reaction (PET-PCR).^14^ Briefly, DNA was extracted from 100 μL of whole blood samples using the QIAamp DNA Mini Kit (Qiagen Inc., Chatsworth, CA, USA). *Plasmodium* genus-specific PET-PCR was performed in duplicate using 5 μL of DNA. Positive controls for PET-PCR consisted of samples with cultured 3D7 Pf or the plasmid for Pv (R64, kindly donated by the Centers for Disease Control and Prevention), and negative control of nuclease-free water were included in each run. Prior to pooling, each clinical sample was characterized for antigen concentration by QPlex. Dilutions of clinical Pf-positive pools were prepared by combining 5 individual Pf-positives, then serially diluting into pooled negative whole blood. Dilutions of clinical Pv-positive pools were prepared by combining 5 individual Pv-positives, then serially diluting into pooled negative whole blood. Additionally, 10 individual Pf-positive and 10 Pv-positive samples were each serially diluted to prepare panels.

### Antigen quantification

All panel members were analyzed for malaria antigen concentration to simultaneously quantify malaria proteins HRP2, PvLDH, pan-malarial LDH, PfLDH, and human C-reactive protein.^15,16^ Each sample was tested both neat and diluted 20-fold to improve the dynamic range of the quantification. Preparation of calibrators and samples and all other steps were conducted according to the protocol. Image collection and analysis were performed using the Q-View Imager Pro and Q-View™ analysis software (Quansys Biosciences).

### Malaria benchmarking panels

A list of the panel members is shown in Table 1. For each panel member, a categorical classification relevant to test performance is listed and the number of serial dilution steps is shown. Most panels target concentrations that span the limits of detection of both conventional and more highly sensitive RDTs. The target analytes relevant to each panel member are also listed.

**Table 1.**
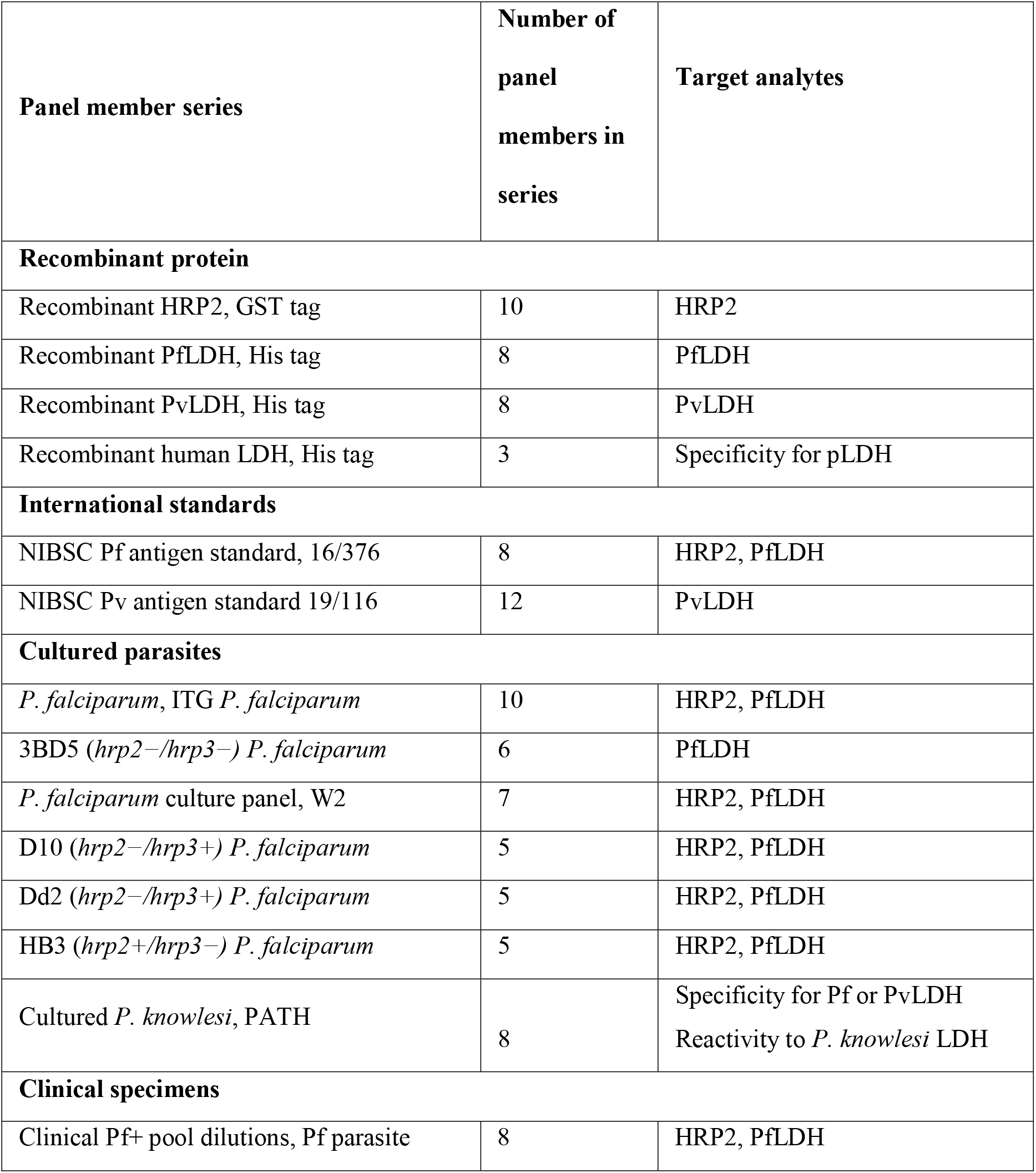
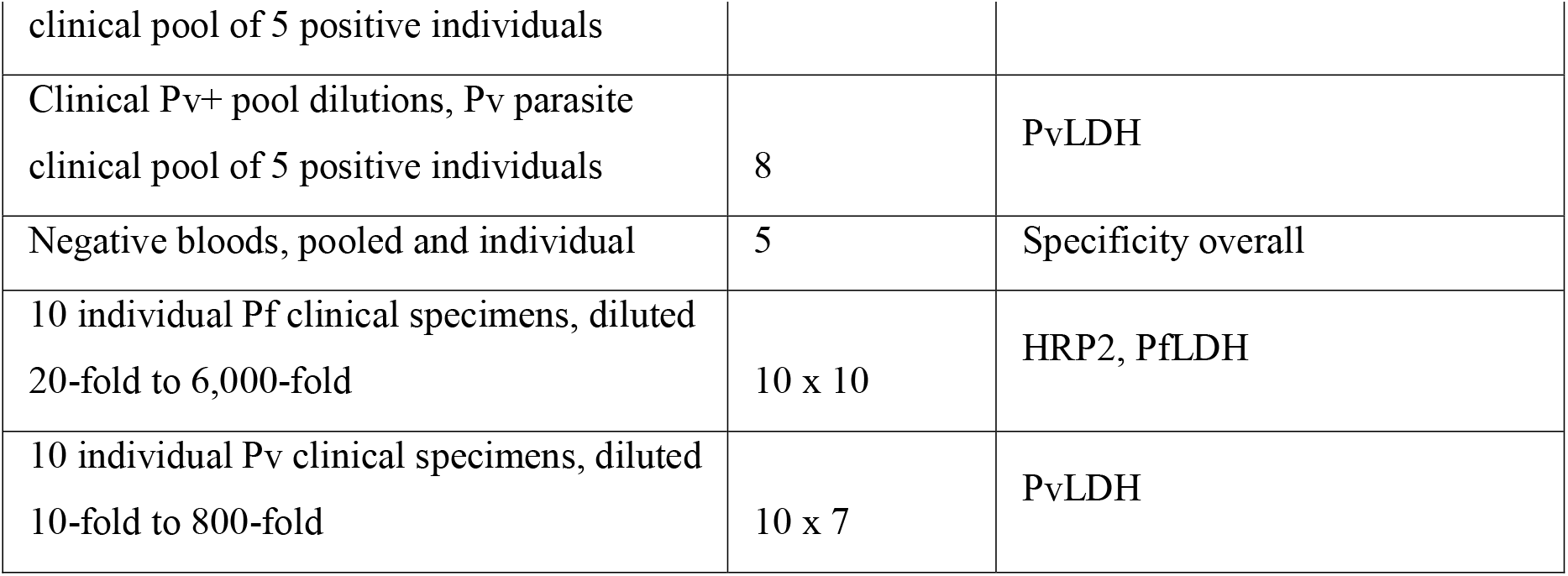
Composition of malaria benchmarking panel. Panel member series descriptions are listed under categories of recombinant protein, international standards, cultured parasites, or clinical specimens, along with the number of dilutions and/or individuals per series. Abbreviations used: GST, glutathione-S-transferase; HRP2, histadine-rich protein 2; PfLDH, *Plasmodium falciparum* lactate dehydrogenase; PvLDH, *Plasmodium vivax* lactate dehydrogenase; LDH, lactate dehydrogenase; pLDH, *Plasmodium* lactate dehydrogenase.

### Tests used in evaluation

Tests used in the evaluation were from Rapigen Inc. (Suwon, South Korea): Rapigen BIOCREDIT Malaria Ag Pf/Pv (pLDH/pLDH) containing test lines to detect PfLDH and PvLDH, and the Rapigen BIOCREDIT Malaria Ag Pf (pLDH/HRPII) containing test lines to detect Pf HRP2 and PfLDH. Comparators for Pf and Pv infection were two best-in-class WHO-prequalified tests containing HRP2 and PfLDH detection lines and HRP2 and PvLDH detection lines.

### Evaluation of RDTs using malaria benchmarking panels

Benchmarking panel member aliquots were kept frozen at −80°C and thawed for up to 2 hours before use and not refrozen or reused after testing. Blood samples were added to tests by calibrated pipettor, in the volume recommended in instructions for use (5 μL). Each test type was run with panel members using three to five replicates for all panel proteins used in testing specificity, and for decreasing concentrations of positive panel members that had adjacent positive results until a clear pattern of negativity was reached with lower concentrations of two or more adjacent dilutions testing negative for 100% of replicates. Concentrations near detection limit were chosen for specific panel members and run with a total of 40 replicate tests above, at, and below concentration identified as near limit of detection. Exceptions to this increased resolution in testing were for positives generated with human recombinant LDH, clinical pool dilutions, clinical individual samples, or culture-derived panel members with limited quantities. Test line intensity was assigned based on comparison to an intensity scale card provided by the manufacturer. Any visible test line was considered positive. All test results were interpreted at times according to manufacturer instructions. If a range of time was given, the earliest time point in the range was used. Test line signal intensity was measured by comparison to a manufacturer-provided scale. All invalid tests were recorded. For each test run, qualitative observations of problems with flow quality, line quality, high background, or test readability were noted.

### Clinical samples used in performance modeling

Human specimens, from which reference data were used in this study, were received from a commercial supplier Discovery Life Sciences (Huntsville, Alabama, USA), from Foundation for Innovative New Diagnostics (FIND, Geneva, Switzerland) Specimen Bank, or collected in malaria research studies. Approvals for these studies for collection of specimens and ongoing malaria research were granted by the following Institutional Review Boards (IRBs), with identification number listed in parentheses following: QIMR Berghofer Medical Research Institute, Brisbane, Australia, Human Research Ethics Committee (2080, 2092, 2098, and 2142, and ACTRN12617000048381); Oxford Tropical Research Ethics Committee, OxTREC, Tak Community Advisory Board, Myanmar, and by the relevant village committee (516-17, 1017-13 and 1015-13); Mali Faculté de Médecine de Pharmacie et d’Odonto Stomatologie, Bamako, Mali and National Institute of Allergy and Infectious Diseases, National Institutes of Health, Bethesda, MD, USA (NCT02334462)); Namibia Ministry of Health and Social Services (17/3/3), and the Institutional Review Boards of the University of Namibia (MRC/259/2017), University of California San Francisco (15–17422) and London School of Hygiene & Tropical Medicine, London, United Kingdom (10411); University of California San Francisco, San Francisco, CA, USA (IRB No.11-05995), Makerere University, Kampala, Uganda (IRB No. 2011-0167); Universidad Peruana Cayetano Heredia (UPCH, Lima, Peru) (UPCH 52707).

Modeling was conducted on a database of reference data from clinical specimens for which the following inclusion criteria were met: (1) available QPlex data for HRP2 and LDH antigen concentrations, (2) positive by either polymerase chain reaction (PCR) or expert microscopy, (3) from natural clinical or asymptomatic infections in humans, and (4) ethics statements indicating IRB approval for sample collection and use. For studies with longitudinal specimen collection, only samples that were positive by microscopy or that were from a pretreatment time point were used to avoid the possibility of persistence of antigen following treatment, which could bias relative concentrations of HRP2 and PfLDH. The final Pf database consisted of 1,001 samples from 9 studies across 12 countries, and the final Pv database consisted of 496 samples from 5 studies across 3 countries.

### Data analysis methods—Estimating the relationship between antigen concentrations and the probability of RDT positivity

For RDTs that only used a single antigen for a given species, a logistic regression model was used with probability of positivity as the independent variable and log10 antigen concentration as the dependent variable. A factor term was also included to account for the sample type (clinical, recombinant, culture, specificity). For RDTs that had two analytes for Pf (i.e., PfLDH/HRP2 tests), two separate logistic regressions models—the same as described above—were run, and then the outputs were combined on a surface to give an estimated probability that either (or both) test lines would be positive based on both HRP2 and PfLDH antigen concentrations. This assumes no cross-reactivity between the two test lines. A Bayesian framework was adopted for all models using the R package brms.^17^ Informative Gaussian priors with a mean of 0 and standard deviation of 3 were used for model coefficients. Each model was run for 10,000 iterations to ensure convergence. Convergence was assessed visually, using a standard convergence diagnostic called R-hat. Statistical analyses were conducted using R 4.2.1 (R Foundation for Statistical Computing, Vienna, Austria). Model predictions and 95% credible intervals were generated using the *fitted* function.

### Data analysis methods—Estimating the impact of the RDTs using clinical sample data

The values at which there was a 90% probability of positivity were extracted from the model for each RDT test line—from hereon these are referred to as the limits of detection. Test limits of detection were then compared to antigen concentrations from the real-world samples and the sensitivity of each test was simulated by calculating the proportion of true positives (RDT-positive and reference-positive) that have antigen concentrations greater than the limit of detection out of all samples positive by the clinical reference data. The samples from the *P. falciparum* database were overlaid on the HRP2 and PfLDH limits of detection and the samples from the *P. vivax* database were overlaid on the PvLDH limits of detection. For RDTs with two analytes for *P. falciparum*, the sensitivity was calculated as the proportion of true positive samples that either had HRP2 concentrations *or* PfLDH concentrations greater than their respective estimated limits of detection.

## RESULTS

### Positivity for Pf detection tests

Replicate test results plotted according to sample concentrations of HRP2 and PfLDH proteins show a clear relationship between RDT positivity and antigen concentration across all benchmark panel members (Figure 1, A and B, and Figure 2, A and B). The logistic regression fits for both HRP2 and PfLDH probability of positivity versus analyte concentration indicate that the Rapigen tests can detect lower concentrations of PfLDH and HRP2 antigen. For PfLDH, the 90% probability of detection was identified at 1,318 pg/mL for the Rapigen Pf (HRPII/pLDH), 525 pg/mL for the Rapigen Pf/Pv, and 5,754 pg/mL for the best-in-class comparator test (Figures 1C, 2C, and 1D). For HRP2, the 90% probability of detection was identified at 525 pg/mL for the Rapigen Pf (HRPII/pLDH), 1,072 pg/mL for the Pf (HRP2/PfLDH) comparator, and 891 pg/mL for the Pf/Pv (HRP2/PvLDH) comparator (Figures 1C, 1D, and 2D). The detection of Pf by combined Pf/Pv tests showed that exclusion of either HRP2, as in the case the Rapigen Pf/Pv test, or exclusion of PfLDH, as with the comparator Pf/Pv test, limits the numbers of samples that are detected overall by HRP2 and/or PfLDH as positive for Pf (Figure 2). A graphical summary of the HRP2 and PfLDH antigen detection limits is shown in Figure 3, and limits of detection with 95% credible intervals are listed in Table 2. The largest gain in sensitivity between the comparator and the Rapigen RDTs is in detection of PfLDH.

**Table 2.** HRP2 and PfLDH antigen concentrations identified at 90% probablity o corresponding 95% credible intervals) for Rapigen and best-in-class WHO-prequalified comparator RDTs.

| Test | Antigen concentration at which test has 90% probability of positivity |  |  |  |
| --- | --- | --- | --- | --- |
|  | HRP2 (pg/mL) |  | PfLDH (pg/mL) |  |
|  | Median estimate | 95% credible interval | Median estimate | 95% credible interval |
| Rapigen Pf (pLDH/HRPII) | 525 | (407–661) | 1318 | (1,175–1,479) |
| Rapigen Pf/Pv (pLDH/pLDH) |  | - | 525 | (447–589) |
| WHO PQ comparator HRP2/PfLDH RDT | 1,072 | (955–1,202) | 5754 | (5,012–6,607) |
| WHO PQ comparator HRP2/PvLDH RDT | 891 | (741–1,047) | - | - |

**Figure 1.**
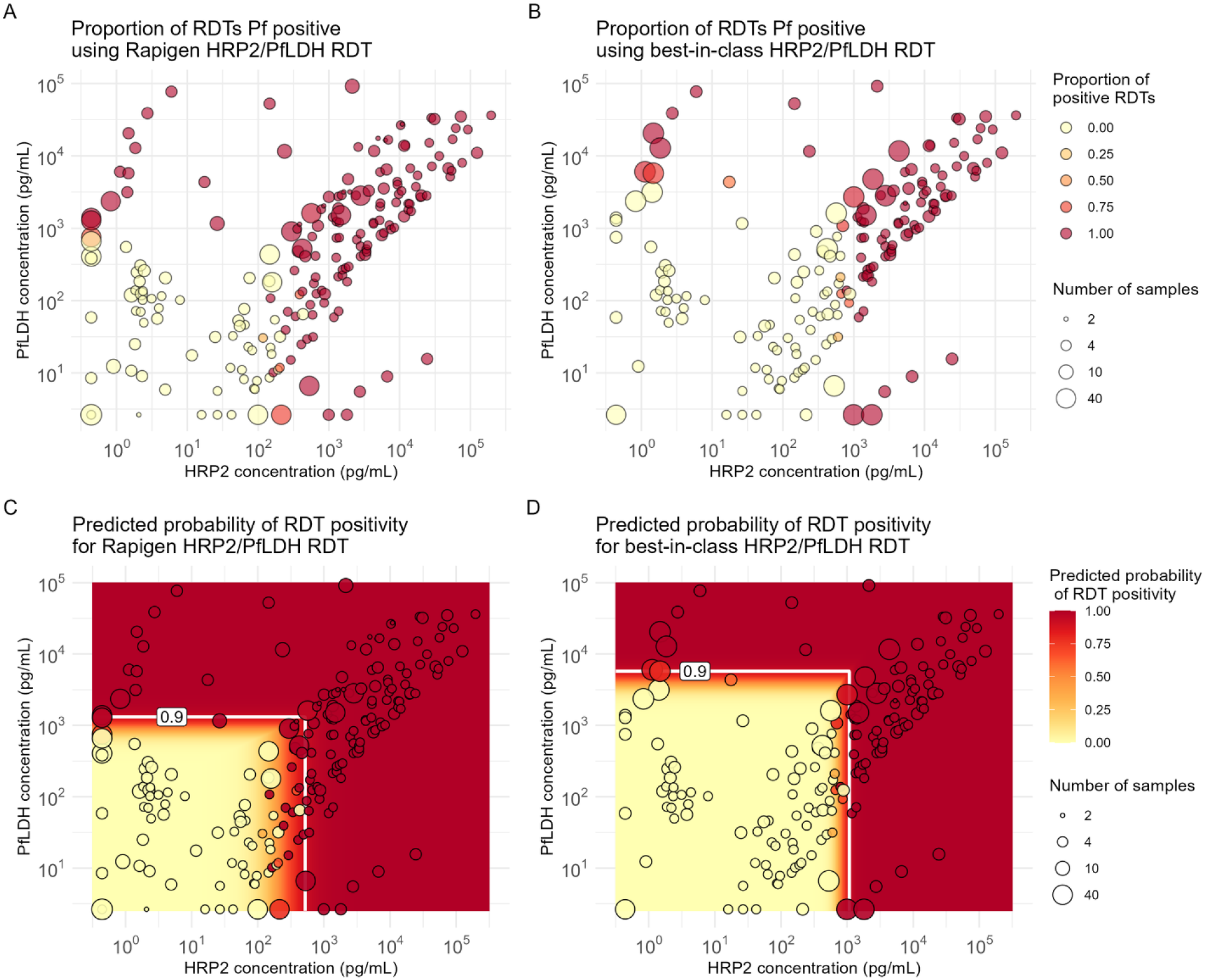
Positivity versus Pf antigen concentrations for Pf detection tests evaluated. A) Proportion of Rapigen BIOCREDIT Malaria Ag Pf (pLDH/HRPII) replicates testing positive, and B) proportion of best-in-class WHO-prequalified comparator Pf test replicates positive and plotted according to HRP2 and PfLDH concentration (pg/mL). Circle size references the number of tests conducted at the concentrations of HRP2 and PfLDH in the sample, indicated by the circle placement. Heat map overlays of C) Figure 1A for Rapigen BIOCREDIT Malaria Ag Pf (pLDH/HRPII) and D) Figure 1B for comparator Pf test, with antigen concentrations at 90% probability of positivity for either PfLDH or HRP2, indicated by white lines.

**Figure 2.**
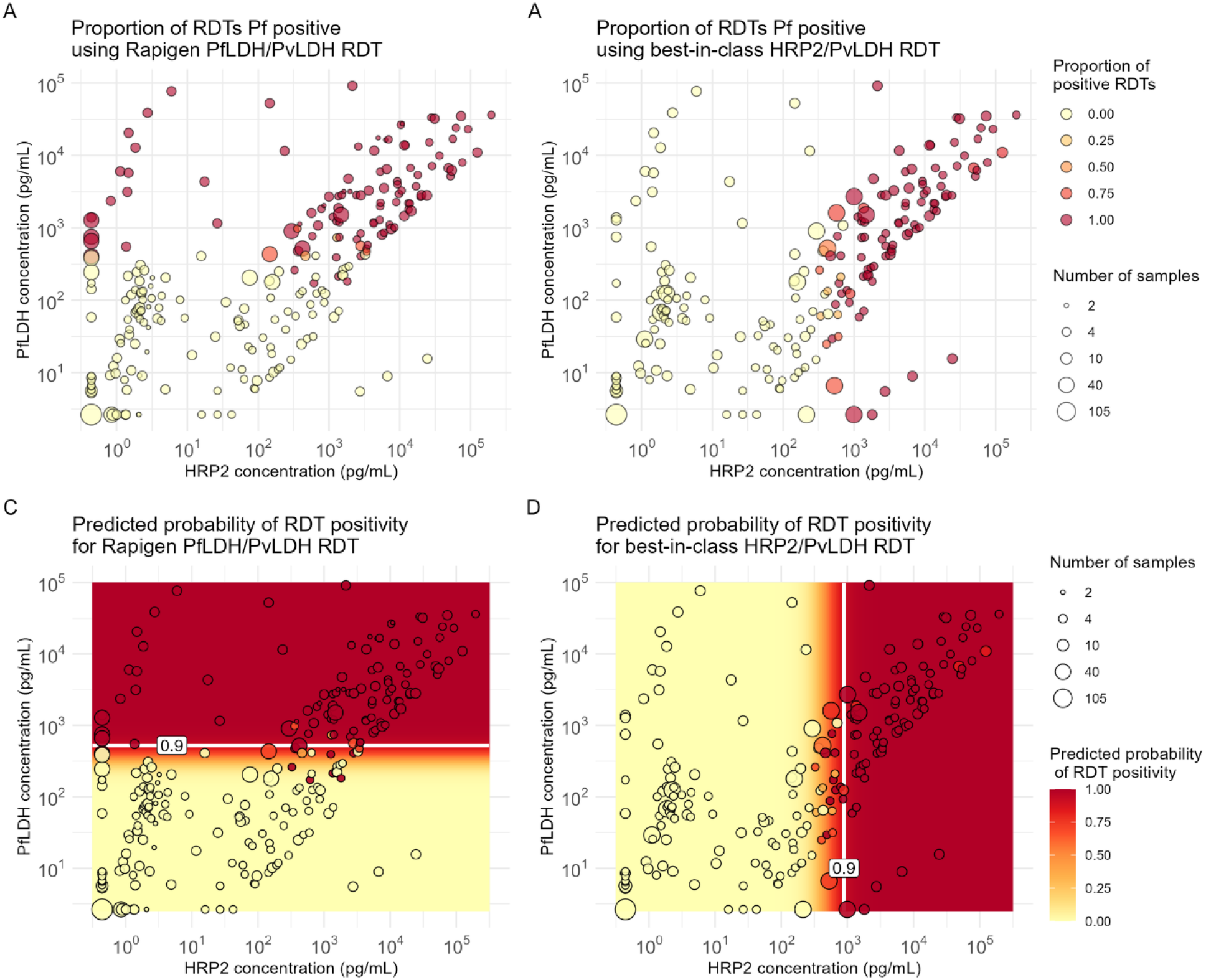
Positivity versus Pf antigen concentrations for Pf/Pv-detection tests evaluated. A) Proportion of Rapigen BIOCREDIT Malaria Ag Pf/Pv (pLDH/pLDH) replicates testing positive, and B) proportion of best-in-class WHO-prequalified comparator Pf/Pv test replicates positive, plotted according to HRP2 and PfLDH concentration (pg/mL). Heat map overlays of C) Figure 2A for Rapigen BIOCREDIT Malaria Ag Pf/Pv (pLDH/pLDH) and D) Figure 2B for comparator Pf/Pv test, with relevant antigen concentrations at 90% probability of positivity, indicated by white lines.

**Figure 3.**
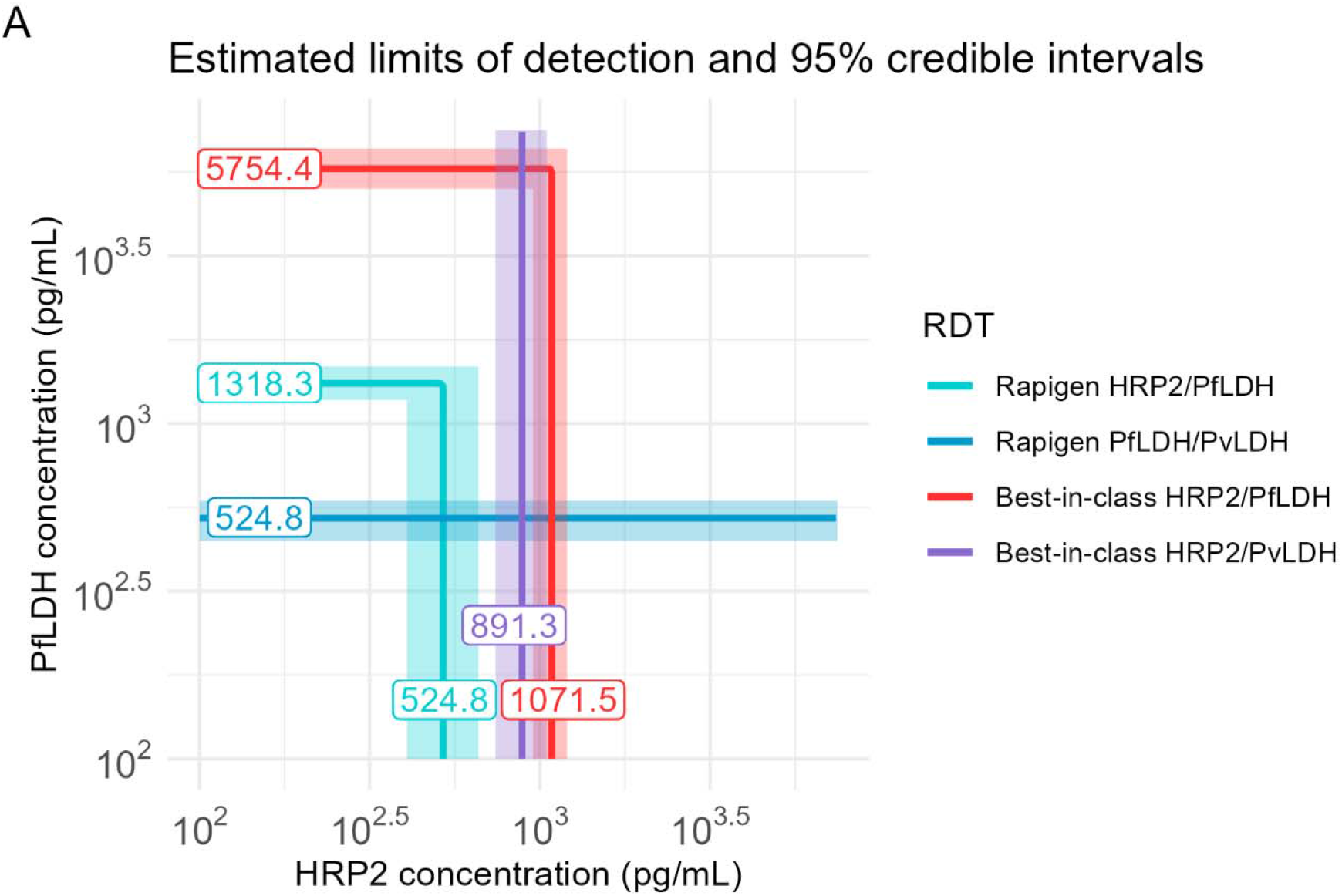
Summary of 90% probability of positivity limits. Results of modeled probability of detection are overlaid for Rapigen Pf (PfLDH/HRP2), Pf/Pv (PfLDH/PvLDH), and best-in-class WHO-prequalified comparator tests for Pf (HRP2/PfLDH) and Pf/Pv (HRP2/PvLDH). Antigen concentrations at the 90% probability, in pg/mL, are indicated on the line.

### Positivity for Pv tests

The Rapigen Pf/Pv (pLDH/pLDH) test can detect lower concentrations of PvLDH in samples, as compared to a best-in-class comparator Pf/Pv test. Modeled positivity versus antigen concentration indicates that the Rapigen Pf/Pv has a 90% probability of detection of 372 pg/mL, and the comparator test 90% probability of detection was found to be at 9,772 pg/mL PvLDH (Figure 4, A and B). Table 3 summarizes the detection limits and corresponding credible intervals. The increase in sensitivity represents an over 10-fold improvement in detection limit with the Rapigen Pf/Pv test as compared to the best-in-class comparator.

**Table 3.** PvLDH antigen concentrations identified at 90% probablity of positivity for Rapigen and best-in-class comparator RDT, with corresponding 95% credible intervals.

| Test | PvLDH antigen concentration (pg/mL) at which test has 90% probability of positivity |  |
| --- | --- | --- |
|  | Median estimate | 95% credible interval |
| Rapigen Pf/Pv (pLDH/pLDH) | 372 | 302–447 |
| Best-in-class HRP2/PvLDH RDT | 9,772 | 7,586–12,883 |

**Figure 4.**
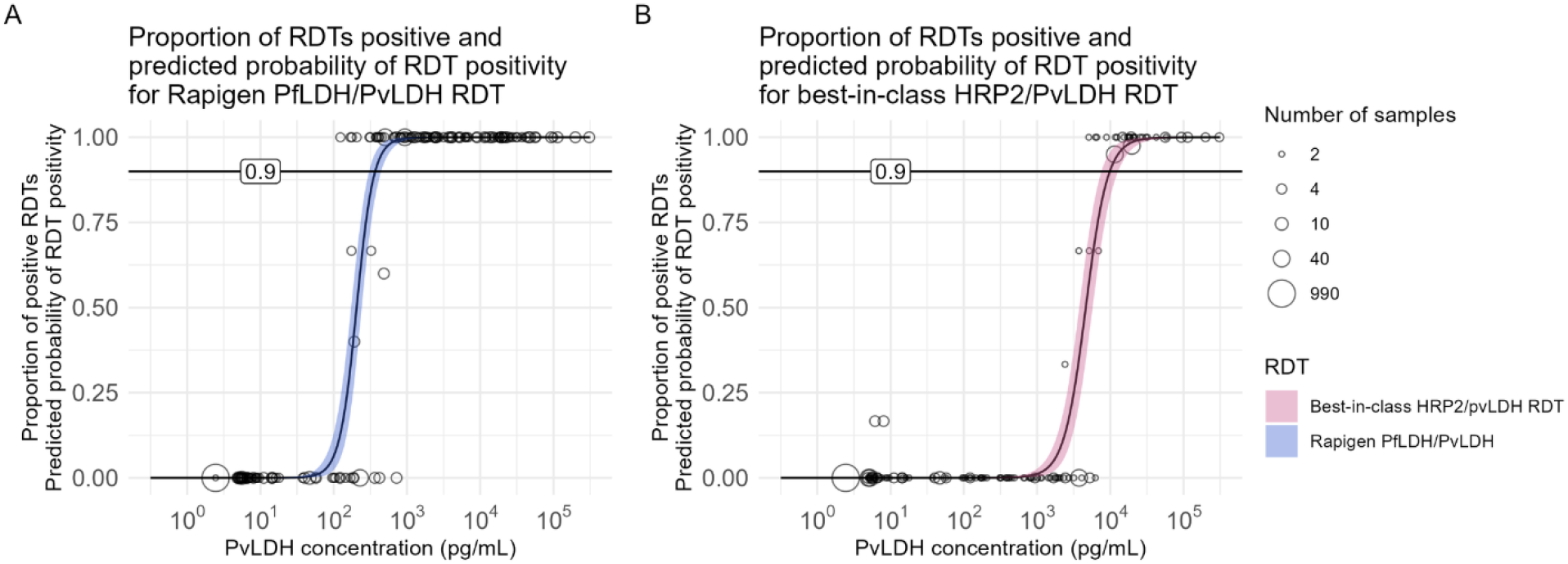
Positivity versus Pv antigen concentrations for Pf/Pv-detection tests evaluated. Proportion of A) Rapigen BIOCREDIT Malaria Ag Pf/Pv (pLDH/pLDH) or B) best-in-class comparator test replicates testing positive for PvLDH versus sample PvLDH concentration (pg/mL). Circle placement indicates the proportion of samples at the given concentration testing positive, and the circle size indicates the number of samples tested at that concentration. The logistic regression fit of probability of positivity for A) Rapigen (blue) and B) best-in-class comparator (pink) is marked with 90% probability.

### Test specificity and reactivity to *P. knowlesi*, and hrp2 or hrp3 gene deletions

All tests and lots used in this evaluation showed high specificity toward malaria-negative donor blood samples and high concentration of human recombinant LDH. Among the strains HB3 (hrp2+/hrp3–), D10, and Dd2 (hrp2–/hrp3+), there were small differences in reactivity in the Rapigen tests, which detected all strains down to 200 parasites/uL or lower, suggesting that the presence of hrp2 or hrp3 individual deletions still provide adequate target analytes for detection at the HRP2 line. The comparator tests preferentially detected the HRP2 over HRP3; D10 was undetectable, and Dd2 was weakly detectable at 2,000 parasites/μL, whereas HB3 was detectable at 200 parasites/μL or lower by the comparators. Cultured *P. knowlesi* was not detected by any test’s PfLDH or HRP2 test lines but was strongly detected by the Rapigen Pf/Pv PvLDH test line down to 125 parasites/μL, whereas the Pf/Pv comparator was only weakly positive at 1,000 parasites/μL.

### Performance of the RDTs on the NIBSC antigen standards

International standards 16/376 for Pf antigen and 19/116 for Pv antigen were tested in concentrations spanning limits of detection of HRP2, PfLDH, and PvLDH of the tests evaluated. As with the other benchmarking panel members, the Rapigen tests showed marked improvements in detectable dilutions compared to the comparator WHO PQ tests for all markers, but especially for PfLDH and PvLDH. Using the Pf standard 16/376, the Rapigen tests were able to detect at least 80% of 5 replicates or at least 90% of 40 replicates run at 3.9 IU/mL HRP2 and 3.9 IU/mL PfLDH (for Rapigen Pf (pLDH/HRPII)), and 1.95 IU/mL PfLDH (for Rapigen Pf/Pv(pLDH/pLDH)). Comparator tests detected at least 80% of 5 replicates or at least 90% of 40 replicates run at 15.6 IU/mL HRP2 (comparator HRP2/PvLDH test) and 15.6 IU/mL HRP2 and 31.3 IU/mL PfLDH (comparator HRP2/PfLDH test).

Using the Pv standard 19/116, the Rapigen Pf/Pv test could detect at least 80% of 5 replicates at 5 IU/mL PvLDH and the comparator could detect at least 80% of 5 replicates at 50 IU/mL PvLDH. Concentrations of target analytes and proportion of positive replicates at all dilutions are shown in Supplemental Material, Tables A and B.

### Estimating malaria detection by the Rapigen and comparator tests using antigen concentration distributions of clinical samples

The performance of the RDTs on clinical specimens was modeled by applying the 90% probability of detection concentrations identified in the benchmarking on clinical sample sets from 751 asymptomatic and 250 febrile patients with corresponding antigen concentration data. While the distributions of HRP2 concentration for asymptomatic and febrile Pf cases strongly overlap, asymptomatic cases reach much lower HRP2 levels. A small peak at low HRP2 concentrations (under limit of quantification of assay) in the HRP2 concentration distribution signals the presence of 32 febrile and 2 asymptomatic HRP2-deleted samples (Figure 5A). The concentration of PfLDH is in general much higher for febrile cases than for asymptomatic ones, though also with large region of overlap. The Rapigen Pf (pLDH/HRPII) RDTs showed only a modest improvement in predicted sensitivity as compared to the comparators for febrile cases and showed a larger improvement for detection of asymptomatic cases. The detection of Pf was found to be the best when both HRP2 and PfLDH were utilized; tests using only PfLDH detection had poorer sensitivity to the comparators, with the exception of when febrile cases with hrp2-deletions are included and performance is compared only to HRP2 detection alone.

**Figure 5.**
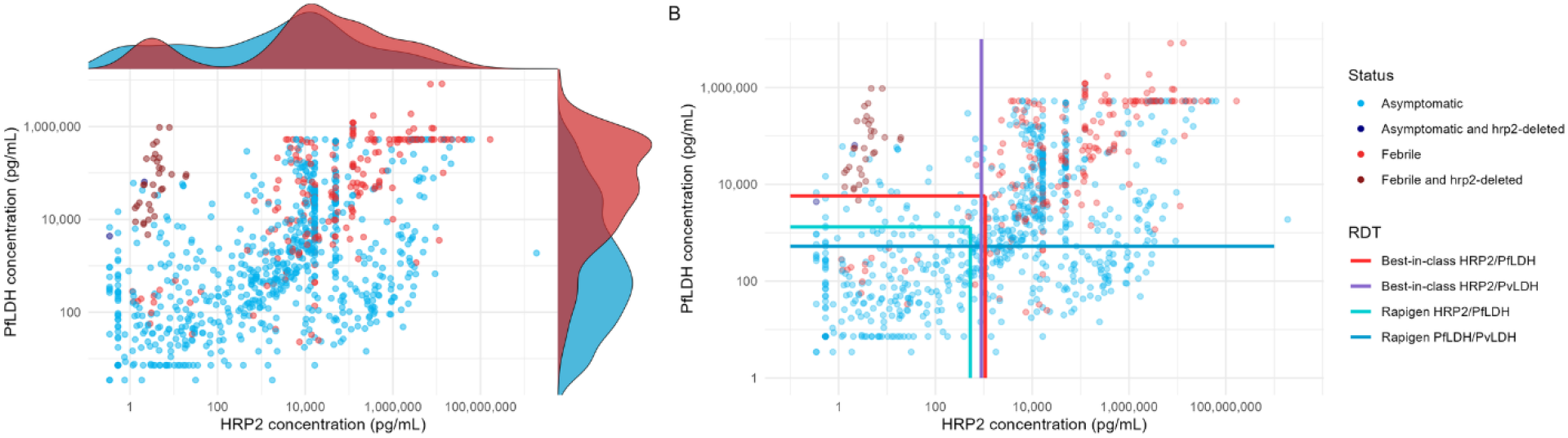
Distribution of antigen concentration in asymptomatic and febrile samples positive for *Plasmodium falciparum* and predicted detection by Rapigen and comparator RDTs. A) PfLDH and HRP2 concentrations are plotted for clinical samples from both symptomatic (dark red, pink) and asymptomatic (dark blue, cyan) patients positive for Pf infection by PCR. Points with either dark red or dark blue coloring represent samples with high probability or confirmed hrp2 gene deletion. Above the plotted values and relative to x-axis HRP2 concentrations, curves show the HRP2 antigen distribution from febrile (red) and asymptomatic (cyan) patients. To the right of the plotted values and relative to the y-axis PfLDH concentration, curves show the PfLDH antigen distribution from febrile (red) and asymptomatic (cyan) patients. B) Plotted points from panel A with overlaid lines marking 90% probability of detection concentrations for Rapigen tests (light and dark turquoise lines) and best-in-class comparator tests (red and purple lines).

**Figure 6.**
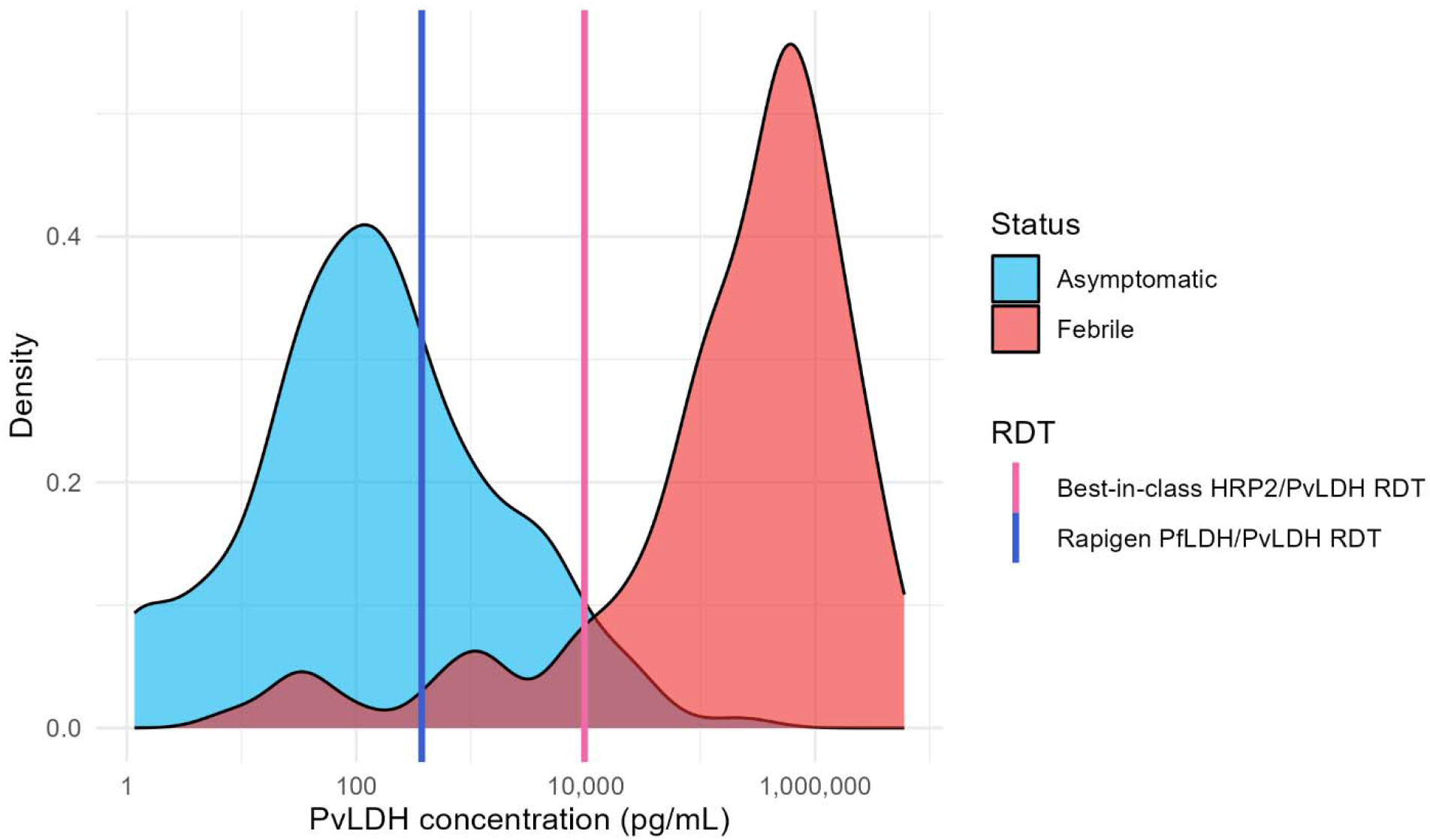
Distribution of antigen concentration in asymptomatic and febrile samples positive for *Plasmodium vivax* and predicted detection by Rapigen and comparator RDTs. A) PvLDH antigen distributions are plotted for clinical samples from both symptomatic (red) and asymptomatic (cyan) patients positive for Pv infection by PCR. Lines marking PvLDH concentrations of 90% probability of detection are overlaid for Rapigen (blue) and best-in-class (pink) tests.

The distribution of PvLDH in febrile and asymptomatic individuals with Pv infection showed a similar pattern of increased antigen levels with febrile cases, but with less overlap between the two groups. Consequently, relative increase in sensitivity of the Rapigen Pf/Pv to the comparator was modest with febrile patients (0.955 vs. 0.881), but there was a large increase in the number of asymptomatic samples with antigen levels that would be detectable, given that very few were predicted to be detected by the comparator (0.326 vs. 0.044).

## Discussion

A benchmarking panel composed of frozen aliquots of recombinant antigens, NIBSC standards, cultured parasites, and clinical specimens was developed to evaluate the performance of RDTs for *P. falciparum* and *P. vivax*. The use of frozen aliquots allows direct comparison of performance in a laboratory setting, with control over variables such as temperature, humidity, sample volume, and user interpretation. While such variables are important factors in clinical settings, reducing variables for performance testing focuses the evaluation on the inherent test performance. The panel members are characterized for concentration of the target antigens relevant to the tests, and inclusion of pLDH recombinant proteins and cultured hrp2/hrp3-deletion mutants allows the interrogation of performance in the absence of HRP2 protein, allowing a better understanding of test performance both in situations where hrp2 and or hrp3 gene deletions may be increasingly prevalent or in high-transmission settings at risk of having persistent HRP2 in circulation following parasite clearance. The relationship between antigen concentration and parasite density is stronger for pLDH than for HRP2 due to its persistence following parasite clearance.^15^ The panel includes NIBSC standards allowing the interpretation of the analytical sensitivity in terms of international units for cross-platform comparisons. Cultured strains can directly assess the reactivity toward strains with hrp2/hrp3 mutations and toward *P. knowlesi*, which has highly variable reactivities among different RDTs and may be important for speciation in coendemic regions.^18–22^ The Rapigen PvLDH test line was found to react in a dose-dependent manner down to 125 parasites/μL, supportive of previous observations of its reactivity with *P. knowlesi*,^23^ whereas the comparator had very little reactivity. The inclusion of a broad range of antigen sources acknowledges that immunoassays may have differing reactivities toward different sources of proteins because there may be small sequence-based, conformational, or even stability differences affecting the epitopes. While limits of detection presented were for the reactivity toward each analyte across the panel members, the limit of detection can also be stratified by source of antigen (i.e., recombinant, culture) to determine if any reactivity bias was present for specific panel components.

Included in the benchmarking panel were the First WHO International Standard for *Plasmodium falciparum* antigen (NIBSC Code: 16/376) and *Plasmodium vivax* antigen (NIBSC Code: 19/116).^24,25^ These standards offer the ability to present RDT limits of detection in standardized international units. There is an opportunity to establish target dilutions or target minimum IU/mL that manufacturers must demonstrate to regulators and procurers as part of their performance criteria. A limitation of the current standards is that they cannot discriminate the contributions of HRP2 and PfLDH concentrations to Pf positivity for RDTs with lines targeting HRP2 combined with other antigens such as PfLDH. This is particularly relevant in understanding the performance of an RDT in the context of hrp2/hrp3-deletions.

It is possible to associate the benchmarking results to performance in clinical settings through the availability of datasets with antigen concentration. Based on the improved detection of pLDH antigen of Pf and Pv by the Rapigen tests, a small improvement in clinical case detection is expected where parasite densities and antigen concentrations tend to be higher. A larger relative improvement to identify subclinical cases is also expected, which will be important as regions move toward elimination or for surveillance programs. While predicted as part of impact modeling for this study, clinical study results have indicated higher sensitivity for the Rapigen tests as compared to standard tests, particularly for subclinical infections.^8^ The improved sensitivity to PfLDH is extremely important to support detection of hrp2/hrp3-deletions. However, in RDTs that only have a PfLDH test line and do not detect HRP2, it is expected to reduce overall sensitivity against HRP2-containing Pf infections. Detection of PfLDH has performed satisfactorily against higher-density panels.^26^

The selection of a broad set of samples for impact modeling may diffuse the antigen distribution patterns that may be found between different geographic regions, degrees of endemicity, and types of study, with the goal to look at overall antigenemia patterns. The distributions of the antigen concentrations showed a large overlap between febrile and asymptomatic cases, but with a clear trend of higher antigen concentrations associated with most febrile cases, except as tempered by inclusion of samples with hrp2 deletion for HRP2 concentrations. The predicted sensitivity toward febrile versus asymptomatic cases was reflective of the trend. To further explore the predicted sensitivity toward samples with higher and lower parasite density, the sensitivities were calculated against samples with equal or less than, and greater than, 200 parasites per μL using the matched sample antigen data to define whether or not it would be detectable by RDT. The antigen distributions parsed by parasite density had a larger overlap than the distributions based on clinical symptoms and correspondingly had less differentiation between the relative sensitivities. However, some of the samples, while PCR-positive, did not have specific parasite density information available and so this analysis is limited by proportions of the sample set that were not classified by density (Supplemental Figures A, Table 2, and Figure B).

Whether the sample sets were stratified by clinical symptoms or parasite density, the BIOCREDIT Malaria Ag Pf (pLDH/HRPII) had a higher predicted sensitivity relative to the comparator and the performance of the BIOCREDIT Malaria Ag Pf/Pv (pLDH/pLDH) was dependent on the presence of hrp2 deletions. When the 34 hrp2-deletion mutant samples were included, the pLDH-only detection was predicted to have poorer sensitivity against the whole sample set relative to a comparator that could detect HRP2 and PfLDH but had superior predicted performance relative to the comparator, which relied solely on HRP2 for Pf detection, thus missing the hrp2-deletion strains (Table 4).

**Table 4.** Modeled clinical performance of RDTs based on analytical detection limits and antigen concentrations of Pf-positive clinical samples. Rapigen and best-in-class comparator RDT 90% probability of detection limits for antigens were compared to antigen concentrations in clinical samples shown in Figure 5 and predicted sensitivity was calculated. Samples were stratified by febrile and asymptomatic infection and results were calculated both without and with (in rightmost column) hrp2 gene deletions included.

| Test name | Febrile status | Including only hrp2-present samples |  | Including all samples |  |
| --- | --- | --- | --- | --- | --- |
|  |  | n | Predicted sensitivity and 95% confidence interval | n | Predicted sensitivity and 95% confidence interval |
| Rapigen Pf (pLDH/HRPII) | Febrile | 218 | 0.936 (0.895, 0.964) | 250 | 0.944 (0.908, 0.969) |
| Rapigen Pf/Pv (pLDH/pLDH) | Febrile | 218 | 0.885 (0.835, 0.924) | 250 | 0.900 (0.856, 0.934) |
| Best-in-class HRP2/PfLDH | Febrile | 218 | 0.927 (0.884, 0.957) | 250 | 0.932 (0.893, 0.96) |
| Best-in-class HRP2/PvLDH | Febrile | 218 | 0.922 (0.878, 0.954) | 250 | 0.804 (0.749, 0.851) |
| Rapigen Pf (pLDH/HRPII) | Asymptomatic | 749 | 0.705 (0.671, 0.737) | 751 | 0.706 (0.672, 0.738) |
| Rapigen Pf/Pv (pLDH/pLDH) | Asymptomatic | 749 | 0.549 (0.512, 0.585) | 751 | 0.55 (0.514, 0.586) |
| Best-in-class HRP2/PfLDH | Asymptomatic | 749 | 0.633 (0.597, 0.667) | 751 | 0.632 (0.597, 0.667) |
| Best-in-class HRP2/PvLDH | Asymptomatic | 749 | 0.611 (0.576, 0.647) | 751 | 0.610 (0.573, 0.645) |

**Table 5.** Modeled clinical performance of RDTs based on analytical detection limits and antigen concentrations of Pv-positive clinical samples. Rapigen and best-in-class comparator RDT 90% probability of detection limits for antigens were compared against clinical samples shown in Figure 6 and sensitivity calculated. Samples were stratified by febrile and asymptomatic infection.

| RDT | Febrile status | n | Predicted sensitivity and 95% confidence interval |
| --- | --- | --- | --- |
| Rapigen Pf/Pv (pLDH/pLDH) | Febrile | 134 | 0.955 (0.905, 0.983) |
| Best-in-class HRP2/PvLDH | Febrile | 134 | 0.881 (0.813, 0.930) |
| Rapigen Pf/Pv (pLDH/pLDH) | Asymptomatic | 362 | 0.326 (0.278, 0.377) |
| Best-in-class HRP2/PvLDH | Asymptomatic | 362 | 0.044 (0.025, 0.071) |

Overall, in laboratory analysis, the Rapigen tests performed well and had similar usability to and had higher sensitivity for a given analyte than the comparator RDTs, suggesting that they will have improved performances in clinical settings and that this improvement will be highly dependent on antigen concentration distributions among the population tested. Although highly controlled laboratory settings do not robustly test usability, we found the tests to have low rates of invalid results or test defects, and they appeared to be of high quality. While malaria RDTs are simple to use, the technique of operators may impact the overall readability and interpretation of the tests.

### Limitations of the study

The testing and impact modeling relies on data generated with an idealized workflow, from frozen specimens only, and with highly controlled sample volumes and laboratory environment, which may not represent clinical use. Variability in test response outside of the test chemistry and analyte concentration may be introduced at a clinical level and could include variable volume from blood sample collection devices, lot differences, temperature and humidity differences, or operator vision. Such sources of variability can become integrated into clinical study performance by default or by design. While benchmarking panels and methodology are designed to identify detection limits and focus on analytical sensitivity, other important components of test accuracy and quality are not comprehensively addressed in these studies., Notably, target analyte specificity needs to be more comprehensively evaluated with malaria-endemic region population samples and with potentially interfering substances or sample types. Cross-reactivity between analyte test lines could be missed in this benchmarking evaluation, especially with high concentrations of non-target *Plasmodium* antigen that would be associated with hyperparasitemia. Companion clinical studies are essential to address performance with clinical use procedures and challenges, and with a breadth of samples that are not readily replicated in the laboratory setting. Given the high degree of sequence homology in LDH across all five human malaria species, inclusion of *P. malariae* and *P. ovale* antigen sources in future panels will be necessary. Clinical specificity cannot be fully interpreted using the methodology described to estimate detection based on antigen concentration alone, with the exception of identifying possible HRP2-persistent samples.

However, these are not tested in this study since samples known to be post-treatment were excluded from the analysis described in this paper. Causes for all cases of false positivity cannot be revealed using antigen concentration analysis. Although inclusion of the hrp2-deleted samples showed some changes in the expected performance for the overall set of samples, the numbers were small, and samples with such deletions may not have been representative of distributions of PfLDH in the dataset.

## Conclusions

A method of evaluating the analytical performance of malaria RDTs is described. From this analysis, the resulting limits of detection were further applied to a dataset of clinical antigen distribution data. Both laboratory analytical sensitivity and corresponding predicted sensitivities with clinical samples are higher for the BIOCREDIT Pf and Pf/Pv RDTs from Rapigen as compared to best-in-class RDT comparators. The increase in sensitivity of the Rapigen RDTs to PfLDH is likely to address to a certain extent the need for RDTs to improve sensitivity even in the presence of hrp2 deletions. However, inclusion of PfLDH detection should still be in combination with detection of HRP2, which offers superior sensitivity relative to PfLDH for Pf.

## Data Availability

All data produced in the present work are contained in the manuscript

## Supplementary material

Supplementary Tables A and B (NIBSC dilutions results)

## Acknowledgments

We thank FIND and study partners for providing sample reference data for clinical antigen concentration analysis that was included in this study. We thank Sharol Hofstedt for assistance in review and editing of the manuscript.

## Authors’ addresses

PATH, 2201 Westlake Ave, Suite 200, Seattle, WA 98121

