## Supplemental Tables A and B for "Analytical sensitivity analysis and clinical impact modeling of Rapigen rapid diagnostics tests for malaria"

Supplemental Materials

Table A. Proportion positive replicate tests at different dilutions of NIBSC 16/376 International standard for Pf antigen. Proportion positive of 5 or 40 replicates shown.

| Control ID | IU/mL of HRP2 and PfLDH | HRP2, pg/mL | PfLDH, pg/mL | Rapigen Pf(pLDH/HRPII) | | Rapigen Pf/Pv | WHO comparator Pf(HRP2/PfLDH) | | WHO comparator Pf/Pv |
|---|---|---|---|---|---|---|---|---|---|
| | | | | HRP2 line | pfLDH line | pfLDH line | HRP2 line | PfLDH line | HRP2 line |
| 16/376 | 62.5 | 4353 | 11661 | 1 | 1 | 1 | 1 | 1 | 1 |
| 16/376 | 31.25 | 1873 | 4760 | 1 | 1 | 1 | 0.98 | 0.73 | 1 |
| 16/376 | 15.63 | 1001 | 2702 | 1 | 1 | 1 | 0.9 | 0 | 1 |
| 16/376 | 7.81 | 566 | 1610 | 0.95* | 1 | 1 | 0 | 0 | 0.78 |
| 16/376 | 3.91 | 295 | 902 | 1 | 1 | 1 | 0 | 0 | 0 |
| 16/376 | 1.95 | 146 | 433 | 0 | 0 | 0.7 | 0 | 0 | 0 |
| 16/376 | 0.98 | 76 | 205 | 0 | 0 | 0 | 0 | 0 | 0 |
| 16/376 | 0.49 | 35 | 94 | 0 | 0 | 0 | 0 | 0 | 0 |
| | *2 replicates had high background and test result couldn't be determined. | | | | | | | | |

Table B. Proportion positive replicate tests at different dilutions of NIBSC 19/116 International Standard for Pv antigen.

| Control | IU/mL | PvLDH, pg/mL | Proportion positive of 5 replicates | |
|---|---|---|---|---|
| | | | Rapigen Pf/Pv | WHO comparator Pf/Pv |
| 19/116 | 400 | 99553 | 1 | 1 |
| 19/116 | 200 | 45480 | 1 | 1 |
| 19/116 | 100 | 23196 | 1 | 1 |
| 19/116 | 50 | 13506 | 1 | 1 |
| 19/116 | 25 | 4122 | 1 | 0.6 |
| 19/116 | 15 | 2534 | 1 | 0 |
| 19/116 | 10 | 1807 | 1 | 0 |
| 19/116 | 5 | 927 | 1 | 0 |
| 19/116 | 2.5 | 484 | 0.6 | 0 |
| 19/116 | 1 | 191 | 0.4 | 0 |
| 19/116 | 0 | 0 | 0 | 0 |
